# Limited cross-variant immune response from SARS-CoV-2 Omicron BA.2 in naïve but not previously infected outpatients

**DOI:** 10.1101/2022.04.07.22273565

**Authors:** Hye Kyung Lee, Ludwig Knabl, Mary Walter, Priscilla A. Furth, Lothar Hennighausen

**Affiliations:** National Institute of Diabetes, Digestive and Kidney Diseases, National Institutes of Health, Bethesda, MD 20892, USA; TyrolPath, Obrist-Brunhuber GmbH, Zams, Austria; Clinical Core, National Institute of Diabetes, Digestive and Kidney Diseases, Bethesda, MD 20892, USA; Departments of Oncology & Medicine, Georgetown University, Washington, DC, USA

## Abstract

Omicron is currently the dominant SARS-CoV-2 variant and several sublineages have emerged. Questions remain about the impact of previous SARS-CoV-2 exposure on cross-variant immune responses elicited by BA.2 infection compared to BA.1. Here we show that without previous history of COVID-19, BA.2 infection induces a reduced immune response against all variants of concern (VOC) compared to BA.1 infection. The absence of ACE2 binding in sera of previously naïve BA.1 and BA.2 patients indicates a lack of meaningful neutralization. In contrast, anti-spike antibody levels and neutralizing activity greatly increased in the BA.1 and BA.2 patients with a previous history of COVID-19. Transcriptome analyses of peripheral immune cells showed significant differences in immune response and specific antibody generation between BA.1 and BA.2 patients as well as significant differences in expression of specific immune genes. In summary, prior infection status significantly impacts the innate and adaptive immune response against VOC following BA.2 infection.

## INTRODUCTION

The highly transmissible SARS-CoV-2 Omicron (B.1.1.529) variant has replaced previous variants and is less susceptible to neutralizing antibodies elicited by vaccination or infection (Cele et al., 2022; Edara et al., 2022; Schmidt et al., 2022). Currently, BA.2 is the dominant Omicron sublineage and has replaced BA.1 (Viana et al., 2022). While BA.1 and BA.2 share 32 mutations, they differ in 28 and demonstrate different antibody neutralization profiles (Arora et al. 2022a). Omicron spike mutations negatively impact neutralizing activity in sera from previously vaccinated and previously infected persons as well as limited efficacy against monoclonal antibody treatment (Dejnirattisai et al., 2022; Hoffmann et al., 2022; Mannar et al., 2022; VanBlargan et al., 2022). Of note, vaccinated individuals with BA.1 infection develop measurable neutralizing antibody titers against BA.1 and BA.2 (Yu et al., 2022). Prior vaccination also broadens the immunological response against variants of concern (VOC) following Omicron infection whereas unvaccinated individuals show an antibody response more limited to the infecting strain (Lee et al., 2022b; Suryawanshi et al., 2022).

The ability of BA.2 infection to induce neutralizing antibodies in unvaccinated individuals, either without or with previous SARS-CoV-2 infection, is pending definition. Here we conducted a comparative investigation of the innate and humoral immune response elicited in 74 Omicron outpatients infected with BA.1 (Lee *et al*., 2022b) or BA.2 that were either naïve prior to Omicron infection or had prior exposure to other SARS-CoV-2 variants. We measured antibody titers and ACE2 binding inhibition as a proxy to neutralization. In addition, we investigated the immune transcriptome and the repertoire of antibody germline alleles in these cohorts.

## RESULTS

### Antibody response after Omicron subvariant infection

We measured antibody titers and neutralizing antibody responses in 74 outpatients infected with either Omicron BA.1 or BA.2 sublineages. Demographic and clinical information are provided in Table 1. While none of the patients had been vaccinated, 26 had been infected by an earlier variant. More than 20% of the BA.1 infected, but none of the BA.2 infected patients, had moderate or severe disease. Humoral immune response was measured between 7 and 21 days (median: days 13-14) post documented Omicron BA.1 or BA.2 infection. In previously uninfected and unvaccinated individuals, anti-Omicron BA.1 spike IgG titers were approximately 40-fold lower following BA.2 infection compared to BA.1 infection (Figure 1A; Table 1). IgG titers against the ancestral spike were almost 70-fold lower following BA.2 as compared to BA.1 infection. IgG antibody titers in individuals previously infected with earlier SARS-CoV-2 variants were at least 5-fold higher with no difference in titers between those infected with the BA.1 versus BA.2 variants (Figure 1A; Table 1). This highlights that previous infection with variants has a particular strong impact on the humoral immune response in BA.2 infected patients.

**Table 1.**
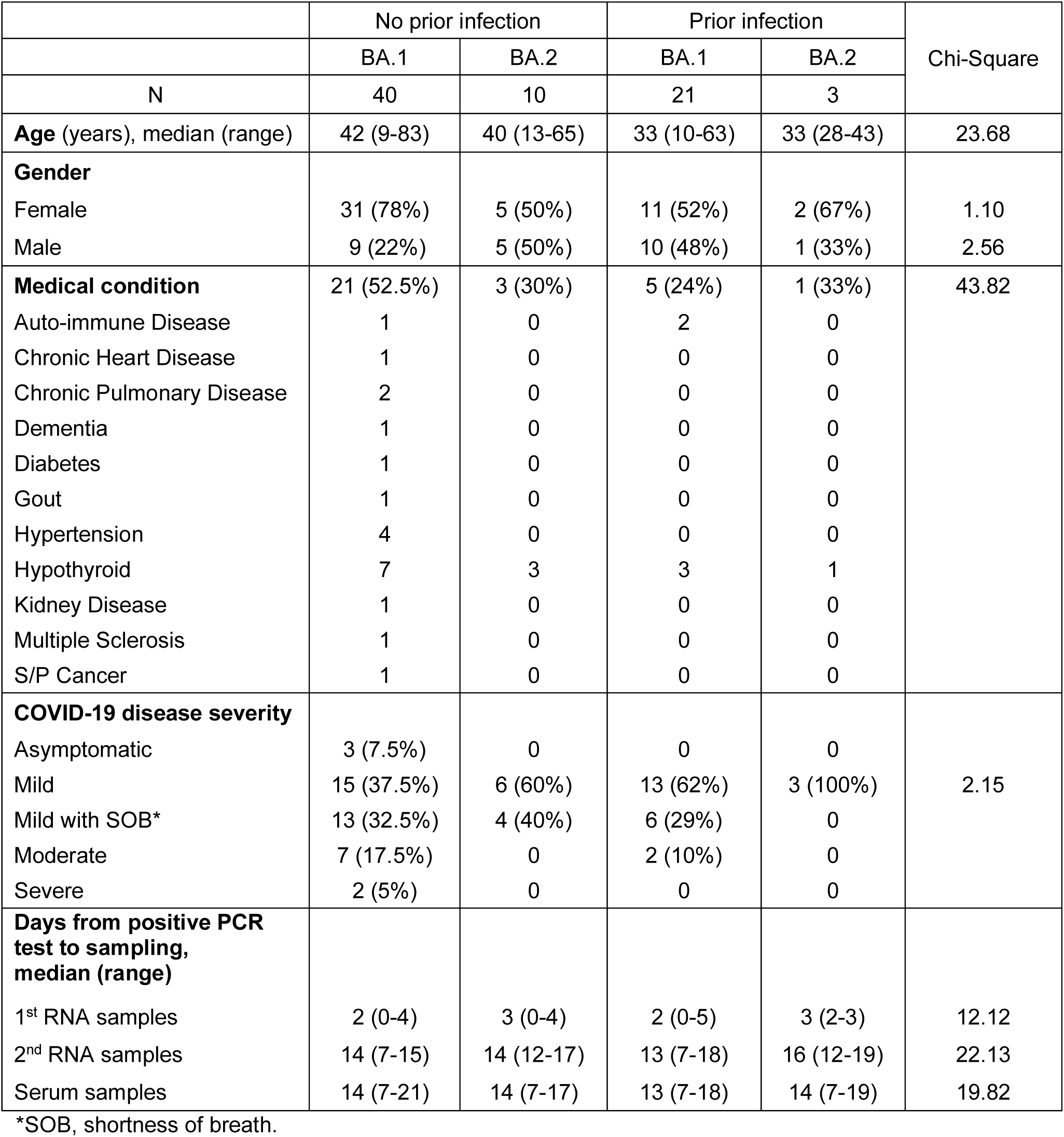
Characteristics of Omicron study population.

**Figure 1.**
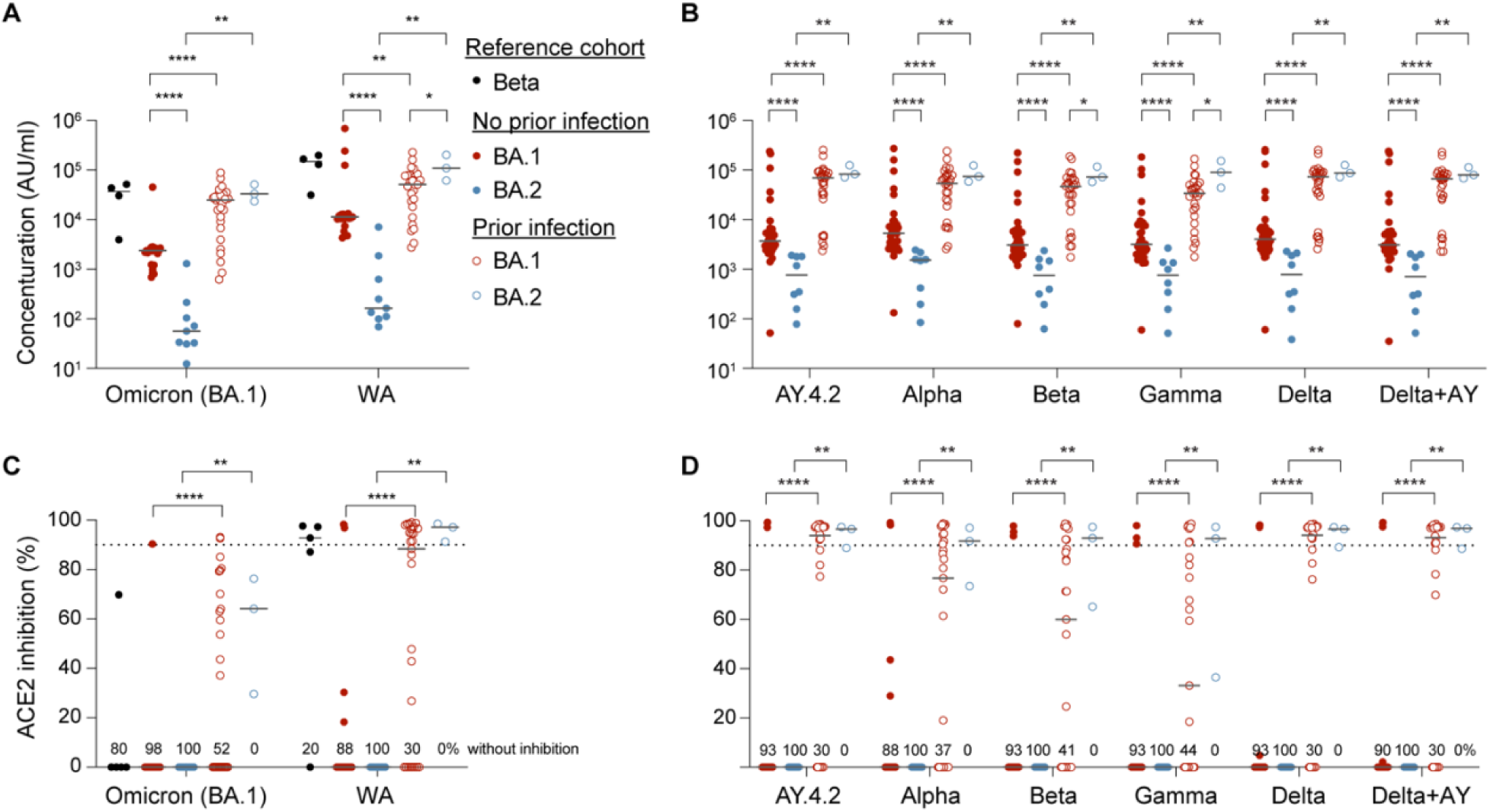
Antibody analysis. (A-B) Plasma IgG antibody binding the SARS-CoV-2 RBD (spike) from the ancestral and Omicron strains (A) as well as the SARS-CoV-2 RBD (spike) from different strains (B) in the Beta patients as a reference, the unvaccinated Omicron patients without or with prior infection experience. WA, authentic SARS-CoV-2 strain. (C-D) Neutralization response to virus spike protein of the ancestral and Omicron variants (A) as well as the SARS-CoV-2 RBD (spike) variants (B). *p*-value between two groups is from one-tailed Mann-Whitney t-test. Percentage of samples with zero neutralization activity for each group are indicated over the X axis. Asterisks indicate significance between groups. \**p* < 0.05, \*\**p* < 0.01, \*\*\**p* < 0.001, \*\*\*\**P* < 0.0001. Line at median, dotted line at 90%.

For reference, titers from individuals infected with the Omicron Beta variant (Knabl et al., 2022) are shown. These approximate those obtained in individuals previously infected with an earlier variant who then experienced BA.1 or BA.2 infection. The differential response in antibody generation following BA.1 and BA.2 infection was consistent across all SARS-CoV-2 variants tested (Figure 1B).

We next measured neutralization capacity using the angiotensin-converting enzyme 2 (ACE2) binding inhibition assay (Figure 1C). Neutralizing activities against the ancestral strain and Omicron BA.1 were similarly low in antigen naïve individuals infected with either BA.1 or BA.2. Similar to overall antibody titers, neutralizing activity was higher in those previously infected. Activity from BA.1 and BA.2 infected individuals with previous SARS-CoV-2 infection approximated that found for individuals infected with the Omicron Beta variant and was similar across all SARS-CoV-2 variants tested against (Figure 1D).

### Transcriptome response

Next, we investigated the impact of BA.1 and BA.2 infection on the immune transcriptome in Omicron patients with or without a previous infection history (Figure 2; Tables S2-3). Buffy coats were isolated within the first three days after validated Omicron infection and bulk RNA-seq was conducted with an average sequencing depth of 200 million reads per sample. First, we directly compared the transcriptomes of the BA.1 and BA.2 infected Omicron cohorts without prior infection. In a direct comparison of the two cohorts, expression of 38 and 15 genes was induced significantly in patients infected with BA.1 and BA.2, respectively (Figure 2A; Table S2). While genes activated upon BA.1 infection are enriched in neutrophil degranulation and innate immune system, the ones in activated upon BA.2 infection are categorized in complement pathway (Figure 2B; Table S2). Genes induced in BA.1 patients, such as C4BPA (Figure 2C), have also been reported to be regulated in COVID-19 patients (Gutmann et al., 2021; Jabeen et al., 2022). Of note, the complement genes C1QA and C1QB that show elevated expression in nonclassical monocytes from COVID-19 patients (Stephenson et al., 2021) are preferentially expressed in BA.2 patients.

**Figure 2.**
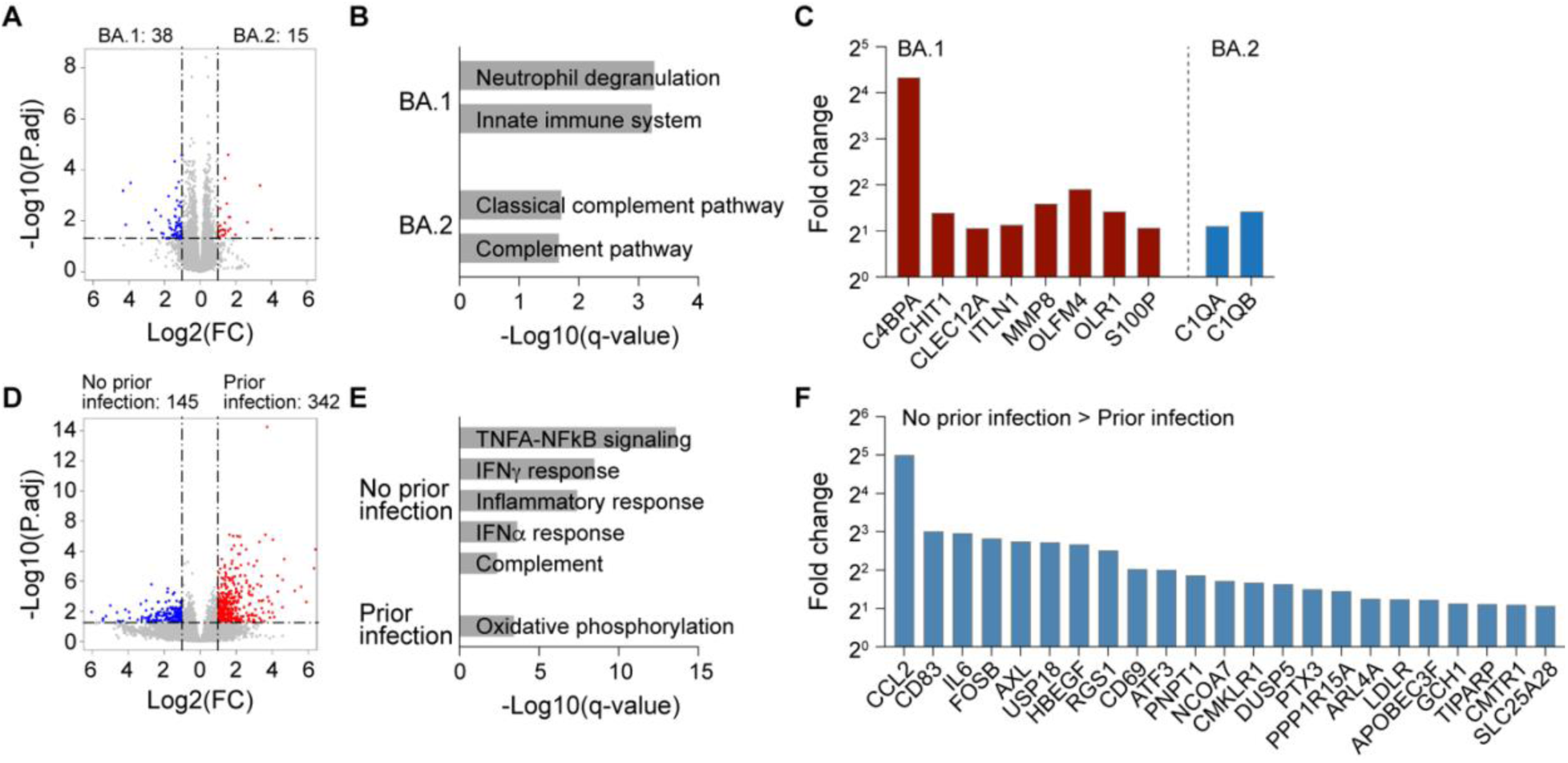
Immune transcriptomes following Omicron infection. (A) Volcano plot of DEGs comparing BA.1 and BA.2 infection in unvaccinated Omicron patients with no prior infection at days 2-3 after infection. (B) Gene categories expressed at significantly higher levels in BA.1 or BA.2 infection group compared to the other. X-axis denotes statistical significance as measured by minus logarithm of FDR q-values. Y-axis ranked the terms by q values. (C) Bar plots with the fold change to mRNA levels between BA.1 and BA.2 groups. Red bars show genes that are higher in the BA.1 group compared to BA.2 and enriched in neutrophil degranulation and innate immune system, and blue bars indicate genes that are higher in the BA.2 group and enriched in complement pathways. (D) Volcano plot of DEGs comparing BA.2 infection in unvaccinated Omicron patients without or with no prior infection at days 2-3 after infection. (E) Gene categories expressed at significantly higher levels in BA.2 infection groups of no prior infection and prior infection. X-axis denotes statistical significance as measured by minus logarithm of FDR q-values. Y-axis ranked the terms by q values. (F) Bar plots with the fold change to mRNA levels in BA.2 groups of no prior infection and prior infection. Blue bars present genes that are higher in the no prior infection group compared to the prior infection group and enriched in innate immune responses.

Next, we compared the immune transcriptomes from BA.2 patients with and without prior infection and identified 342 and 145 significantly induced genes, respectively (Figure 2D; Table S3). GSEA analyses linked the induced genes in the ‘no prior infection’ cohort to innate immune responses, including interferon and inflammatory pathways (Figure 2E). Genes activated in the ‘no prior infection’ cohort (Figure 2F), such as CCL2 and USP18, are part of the interferon response in severe COVID-19 (Cao, 2021; Munnur et al., 2021).

### Antibody germline repertoire

The higher level of anti-spike antibodies and neutralization activity in BA.1 patients of the ‘no prior infection’ group compared to BA.2 infection, led us to dig deeper and interrogate the germline antibody gene usage. Specifically, we determined the range of immunoglobulin heavy chain (IGHV) gene usage elicited by BA.1 and BA.2 sublineage infection in the ‘no previous infection’ cohort (Figure 3). RNA-seq was conducted 13-16 days after the positive PCR test. The number of total immunoglobulin genes (heavy chain variable (IGHV), light chain variable (IGKV and IGLV) and T cell receptor alpha and beta variable (TRAV and TRBV)) revealed the use of a broad range of germlines in all cohorts without statistically significance (Figure 3A). However, we observed increased frequency of transcription of eight VH genes, including IGHV3-11, IGHV3-66, IGHV1-2 and IGHV4-61, in the BA.1 infection group compared to the BA.2 cohort (Figure 3B). These particular heavy chain variants have been identified in SARS-CoV-2 infected patients and COVID-19 vaccinated individuals that developed strongly neutralizing antibodies (Andreano et al., 2021; Andreano and Rappuoli, 2021; Collier et al., 2021; He et al., 2021; Lee et al., 2022a; Zhang et al., 2021). Expression of some of these genes was also activated in octogenarians which had received a single dose of the BNT162b mRNA vaccine 15 months after recovery from COVID-19 (Lee *et al*., 2022a).

**Figure 3.**
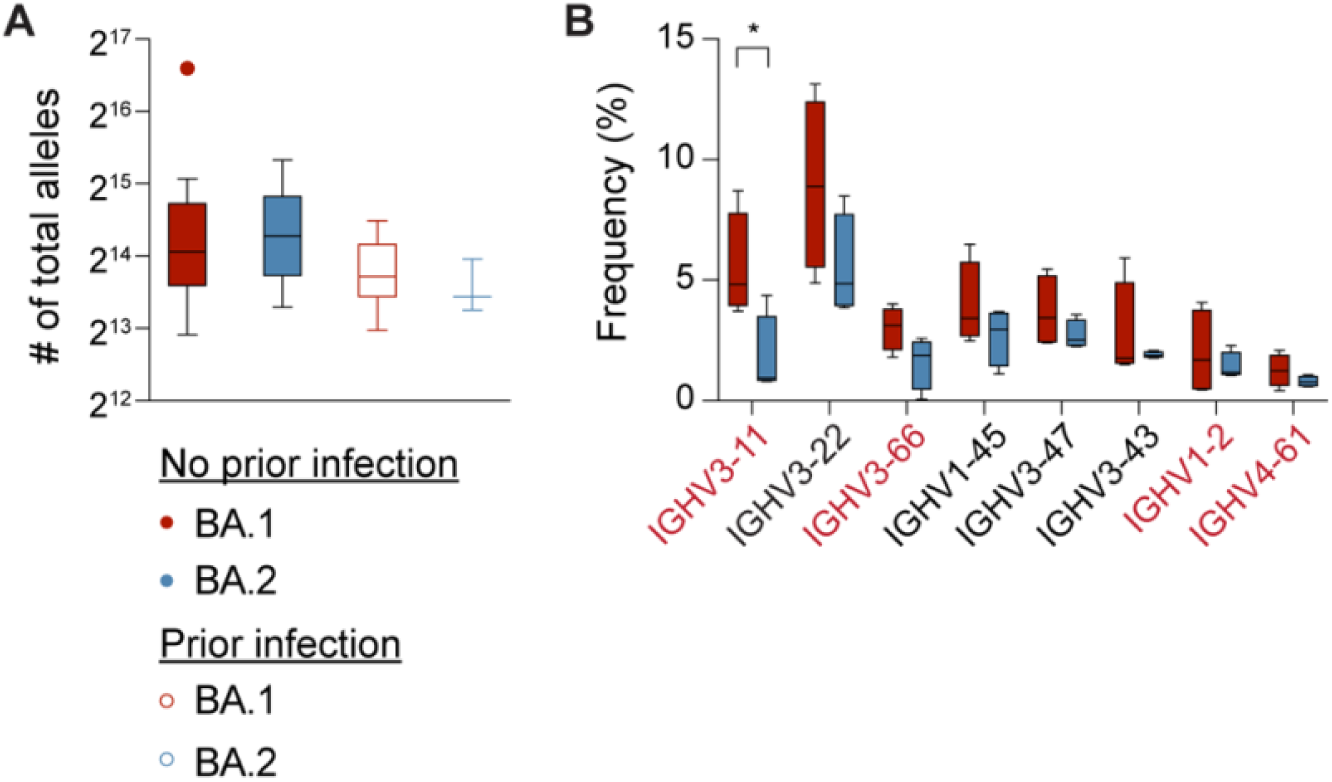
SARS-CoV-2-RBD-specific B-cell memory in Omicron patients. (A) The number of total alleles that are generated at days 13-14. (B) Box plots show the frequency (%) of variable (V) gene usage for all paired heavy chains in BA.1 and BA.2 infection of the no prior infection groups. Ones identified in neutralizing antibodies to COVID-19 infection are marked in red. *p*-value between two groups is from one-tailed Mann-Whitney t-test. \**p* < 0.05. Median, middle bar inside the box; IQR, 50% of the data; whiskers, 1.5 times the IQR.

## DISCUSSION

While the Omicron BA.1 and BA.2 sublineages share mutations they also carry unique mutations, 28 in BA.2, which can explain their differential response to vaccine-induced and therapeutic antibodies (Ai et al., 2022; Arora et al., 2022a; Arora et al., 2022b). Previous studies have also demonstrated that BA.1 infection of naïve persons yields a very limited antibody response to SARS-CoV-2 VOCs other than Omicron itself (Lee *et al*., 2022b; Rössler et al., 2022). However, prior vaccination or prior infection with older SARS-CoV-2 variants augment the immune response upon BA.1 breakthrough infection (Lee *et al*., 2022b; Rössler *et al*., 2022). Our study now reveals that in antigen naïve individuals the immunologic response following BA.2 infection is even lower than that elicited by BA.1 infection as indicated by significantly lower antibody titers against all VOCs. This also highlights that the set of 28 mutations unique to BA.2 contribute to the profound immune evasion.

While other studies have focused on the humoral response including neutralizing activity induced by Omicron infection (Suryawanshi *et al*., 2022), we have extended the investigations and explored the presence of germline immunoglobulin variants induced by BA.1 and BA.2 infection. RBD-targeting antibodies can block SARS-CoV-2 binding to ACE2 and neutralizing immunoglobulin G heavy-chain variable (IGHV) gene usage has been reported (Yuan et al., 2020). Our study identified several IGHV germline alleles in Omicron patients that have been identified in neutralizing antibodies in COVID-19 patients (Yan et al., 2021; Yuan *et al*., 2020).

In conclusion, our study demonstrates that Omicron BA.2 infection of unvaccinated individuals without prior COVID-19 does not elicit effective antibodies against BA.1 and non-Omicron variants. In previously infected individuals, however, BA.2 infection successfully induces immunity against other variants.

### Limitations of the study

There are several limitations to the current study. First, the study was conducted on volunteers from a specific geographical area, Tyrol (Austria). Secondly, limitation is the confinement of the study to a timeframe of three weeks following Omicron infection. Third, the number of the BA.2 patients with the prior infection experiment is limited. Finally, some of the data collected on breakthrough infections was reliant on retrospective chart review and not collected as part of a prospective study.

## Data Availability

RNA-seq data generated from this study will be deposited in the Gene Expression Omnibus (GEO) before publishing the manuscript.
RNA-seq data of Omicron patients were obtained under GSE190747.

## ACKNOWLEDGMENTS

This work was supported by the Intramural Research Program (IRP) of the National Institute of Diabetes and Digestive and Kidney Diseases (NIDDK).

Our gratitude goes to the participants who contributed to this study to advance our understanding of SARS-CoV-2 Omicron infection. We thank Yuhai Dai from the NIDDK clinical core for helping antibody assay. This work was utilized the computational resources of the NIH HPC Biowulf cluster (http://hpc.nih.gov). RNA-sequencing and single cell RNA-sequencing was conducted in the NIH Intramural Sequencing Center, NISC (https://www.nisc.nih.gov/contact.htm)

## AUTHOR CONTRIBUTIONS

H.K.L., L.K. and L.H. designed the study. L.K recruited patients and collected material. H.K.L. analyzed RNA-seq data. M.W. conducted antibody assays. H.K.L., L.K., P.A.F. and L.H. analyzed data. L.H. supervised project. H.K.L., L.K., P.A.F. and L.H. wrote the paper. All authors read and approved the manuscript.

## DECLARATION OF INTERESTS

The authors declare not competing interests.

## INCLUSION AND DIVERSITY

We worked to ensure ethnic or other types of diversity in the recruitment of human subjects. We worked to ensure that the study questionnaires were prepared in an inclusive way. While citing references scientifically relevant for this work, we also actively worked to promote gender balance in our reference list. The author list of this paper includes contributors from the location where the research was conducted who participated in the data collection, design, analysis, and/or interpretation of the work.

## STAR METHODS

### KEY RESOURCES TABLE

| REAGENT or RESOURCE | SOURCE | IDENTIFIER |
| --- | --- | --- |
| Biological Samples |  |  |
| Human serum | Tyrol, Austria | N/A |
| Human immune cells | Tyrol, Austria | N/A |
| Critical Commercial Assays |  |  |
| Maxwell RSC simply RNA Blood Kit | Promega | Cat# ASB1380 |
| V-PLEX SARS-CoV-2 Panel 23 (IgG) Kit | Meso Scale Discovery | Cat# K15567U |
| V-PLEX SARS-CoV-2 Panel 23 (ACE2) Kit | Meso Scale Discovery | Cat# K15570U |
| TruSeq Stranded mRNA Library Prep Kit | Illumina, | Cat# RS-20020595 |
| Deposited Data |  |  |
| Raw and analyzed data | This paper | GEO: GSE |
| Omicron patient (Days 2-3) | Lee et al., 2022b | GEO: GSE190747 |
| Human reference genome UCSC, hg19 | UCSC Genome Browser | http://hgdownload.soe.ucsc.edu/downloads.html#mouse |
| Software and Algorithms |  |  |
| MSD DISCOVERY WORKBENCH analysis software |  | https://www.mesoscale.com/en/products_and_services/software |
| Trimmomatic (version 0.36) | Bolger et al., 2014 | http://www.usadellab.org/cms/?page=trimmomatic |
| STAR (2.5.4a) | Dobin et al., 2013 | https://anaconda.org/bioconda/star/files?version=2.5.4a |
| HTSeq | Anders et al., 2015 | https://htseq.readthedocs.io/en/master/ |
| R (3.6.3) |  | https://www.R-project.org/ |
| Bioconductor | Huber et al., 2015 | https://www.bioconductor.org/ |
| DESeq2 | Love et al., 2014 | https://bioconductor.org/packages/release/bioc/html/DESeq2.html |
| RUVSeq | Risso et al., 2014 | https://bioconductor.org/packages/release/bioc/html/RUVSeq.html |
| dplyr |  | https://CRAN.R-project.org/package=dplyr |
| ggplot2 | Wickham, 2009 | https://ggplot2.tidyverse.org/ |
| GSEA |  | https://www.gsea-msigdb.org/gsea/msigdb |
| MiXCR | Bolotin et al., 2015 | https://mixcr.readthedocs.io/en/master/ |
| PRISM GraphPad (9.0.0) |  | https://www.graphpad.com/scientific-software/prism/ |

## RESOURCE AVAILABILITY

### Lead Contact

Further information and requests for resources and reagents should be directed to and will be fulfilled by the Lead Contact Lothar Hennighausen.

### Materials Availability

This study did not generate new unique reagents.

### Data and Code Availability

- RNA-seq data generated from this study will be deposited in the Gene Expression Omnibus (GEO) before publishing the manuscript.
- RNA-seq data of Omicron patients were obtained under GSE190747.
- This paper does not report original code.
- Any additional information required to reanalyze the data reported in this paper is available from the lead contact upon request.

## EXPERIMENTAL MODEL AND SUBJECT DETAILS

### Study population, study design and recruitment

Data from a total of 74 patients infected with either Omicron BA.1 or BA.2 were analyzed. 13 BA.2 patients and 10 new BA.1 patients were recruited for the study under informed consent 50 with no history of prior vaccination and prior infection with other SARS-CoV-2 variants and 24 unvaccinated patients with prior infections (Table 1). Recruitment and blood sample collection took place between December 2021 and March 2022. Day numbers for samples refers to the number of days following initial positive SARS-CoV-2 RT-PCR test. An additional 53 BA.1 patients from our previous study (Lee *et al*., 2022b) were included. Patient recruitment was performed by a medic who assessed clinical status including performance of an oxygen saturation (SpO2) test. The clinical spectrum of the patients’ SARS-CoV-2 symptoms were classified based on the National Institutes of Health (NIH) treatment guidelines (https://files.covid19treatmentguidelines.nih.gov/guidelines/section/section_43.pdf). This study was approved (EK Nr: 1064/2021) by the Institutional Review Board (IRB) of the Office of Research Oversight/Regulatory Affairs, Medical University of Innsbruck, Austria, which is responsible for all human research studies conducted in the State of Tyrol (Austria). The investigators do not need to have an affiliation with the University of Innsbruck. Written informed consent was obtained from all subjects. Participant information was coded and anonymized. This study was determined to impose minimal risk on participants. All methods were carried out in accordance with relevant guidelines and regulations. All research has been have been performed in accordance with the Declaration of Helsinki (https://www.wma.net/policies-post/wma-declaration-of-helsinki-ethical-principles-for-medical-research-involving-human-subjects/). In addition, we followed the ‘[Sex and Gender Equity in Research – SAGER – guidelines](https://researchintegrityjournal.biomedcentral.com/articles/10.1186/s41073-016-0007-6)’ and included sex and gender considerations where relevant.

## METHOD DETAILS

### Antibody assay

Whole blood was collected by medical personnel after subjects had tested positive for SARS-CoV-2. Antibody containing sera were obtained by centrifuging EDTA blood samples for 10 min at 4,000g. End-point binding IgG antibody titers to various SARS-CoV-2–derived antigens were measured using the Meso Scale Discovery (MSD) platform. SARS-CoV-2 spike, nucleocapsid, Alpha, Beta, Gamma, Delta, and Omicron spike subdomains were assayed using the V-plex multispot COVID-19 serology kits (Panel 23 (IgG) Kit, K15567U). Plates were coated with the specific antigen on spots in the 96 well plate and the bound antibodies in the samples (1:50000 dilution) were then detected by anti-human IgG antibodies conjugated with the MSD SULPHO-TAG which is then read on the MSD instrument which measures the light emitted from the tag.

### ACE2 binding inhibition (Neutralization) ELISA

The V-PLEX COVID-19 ACE2 Neutralization kit (Meso Scale Discovery, Panel 23 (ACE2) Kit, K15570U) was used to quantitatively measure antibodies that block the binding of ACE2 to its cognate ligands (SARS-CoV-2 and variant spike subdomains). Plates were coated with the specific antigen on spots in the 96 well plate and the bound antibodies in the samples (1:10 dilution) were then detected by Human ACE2 protein conjugated with the MSD SULPHO-TAG which is then read on the MSD instrument which measures the light emitted from the tag.

### Extraction of the buffy coat and purification of RNA

Whole blood was collected, and total RNA was extracted from the buffy coat and purified using the Maxwell RSC simply RNA Blood Kit (Promega) according to the manufacturer’s instructions. The concentration and quality of RNA were assessed by an Agilent Bioanalyzer 2100 (Agilent Technologies, CA).

### mRNA sequencing (mRNA-seq) and data analysis

The Poly-A containing mRNA was purified by poly-T oligo hybridization from 1 mg of total RNA and cDNA was synthesized using SuperScript III (Invitrogen, MA). Libraries for sequencing were prepared according to the manufacturer’s instructions with TruSeq Stranded mRNA Library Prep Kit (Illumina, CA, RS-20020595) and paired-end sequencing was done with a NovaSeq 6000 instrument (Illumina) yielding 200-350 million reads per sample.

The raw data were subjected to QC analyses using the FastQC tool (version 0.11.9) (https://www.bioinformatics.babraham.ac.uk/projects/fastqc/). mRNA-seq read quality control was done using Trimmomatic (Bolger et al., 2014) (version 0.36) and STAR RNA-seq (Dobin et al., 2013) (version STAR 2.5.4a) using 150 bp paired-end mode was used to align the reads (hg19). HTSeq (Anders et al., 2015) (version 0.9.1) was to retrieve the raw counts and subsequently, Bioconductor package DESeq2 (Love et al., 2014) in R (https://www.R-project.org/) was used to normalize the counts across samples (Zhao et al., 2021) and perform differential expression gene analysis. Additionally, the RUVSeq (Risso et al., 2014) package was applied to remove confounding factors. The data were pre-filtered keeping only genes with at least ten reads in total. The visualization was done using dplyr (https://CRAN.R-project.org/package=dplyr) and ggplot2 (Wickham, 2009). The genes with log2 fold change >1 or <-1 and adjusted p-value (pAdj) <0.05 corrected for multiple testing using the Benjamini-Hochberg method were considered significant and then conducted gene enrichment analysis (GSEA, https://www.gsea-msigdb.org/gsea/msigdb).

For T- or B-cell receptor repertoire sequencing analysis, trimmed fastq files from bulk RNA-seq were aligned against human V, D and J gene sequences using the default settings with MiXCR (Bolotin et al., 2017; Bolotin et al., 2015).

### Quantification and statistical analysis

Differential expression gene (DEG) identification used Bioconductor package DESeq2 in R. P-values were calculated using a paired, two-side Wilcoxon test and adjusted p-value (pAdj) corrected using the Benjamini–Hochberg method. The cut-off value for the false discovery rate was pAdj > 0.05. Genes with log2 fold change >1 or <-1, pAdj <0.05 and without 0 value from all sample were considered significant. For significance of each GSEA category, significantly regulated gene sets were evaluated with the Kolmogorov-Smirnov statistic. P-values of antibody between two groups were calculated using one-tailed Mann-Whitney t-test on GraphPad Prism software (version 9.0.0). A value of \**P* < 0.05, \*\**P* < 0.01, \*\*\**P* < 0.001, \*\*\*\**P* < 0.0001 was considered statistically significant.

## Notes

### Competing Interest Statement

The authors have declared no competing interest.

### Author Declarations

This study was approved (EK Nr: 1064/2021) by the Institutional Review Board (IRB) of the Office of Research Oversight/Regulatory Affairs, Medical University of Innsbruck, Austria, which is responsible for all human research studies conducted in the State of Tyrol (Austria). Participant information was coded and anonymized.

